# Incorporating the mutational landscape of SARS-COV-2 variants and case-dependent vaccination rates into epidemic models

**DOI:** 10.1101/2021.11.28.21266882

**Authors:** Mohammad Mihrab Chowdhury, Md Rafiul Islam, Md Sakhawat Hossain, Nusrat Tabassum, Angela Peace

## Abstract

Coronavirus Disease (COVID-19), which began as a small outbreak in Wuhan, China in December 2019, became a global pandemic within months due to its high transmissibility. In the absence of pharmaceutical treatment, various non-pharmaceutical interventions (NPIs) to contain the spread of COVID-19 brought the entire world to a halt. After almost a year of seemingly returning to normalcy with the world’s quickest vaccine development, the emergence of more infectious and vaccine resistant coronavirus variants is bringing the situation back to where it was a year ago. In the light of this new situation, we conducted a study to portray the possible scenarios based on the three key factors : impact of interventions (pharmaceutical and NPIs), vaccination rate, and vaccine efficacy. In our study, we assessed two of the most crucial factors, transmissibility and vaccination rate, in order to reduce the spreading of COVID-19 in a simple but effective manner. In order to incorporate the time-varying mutational landscape of COVID-19 variants, we estimated a weighted transmissibility composed of the proportion of existing strains that naturally vary over time. Additionally, we consider time varying vaccination rates based on the number of daily new cases. Our method for calculating the vaccination rate from past active cases is an effective approach in forecasting probable future scenarios as it actively tracks people’s attitudes toward immunization as active cases change. Our simulations show that if a large number of individuals cannot be vaccinated by ensuring high efficacy in a short period of time, adopting NPIs is the best approach to manage disease transmission with the emergence of new vaccine breakthrough and more infectious variants.

## 1. Introduction

The catastrophic effect of the H1N1 virus (Influenza Flu) in the twentieth century has returned with the worldwide spread of coronavirus. Coronavirus disease (COVID-19) is an excellent illustration of what may happen if the proper preventive measures are not taken in a timely way, even in this advanced age of technology and medical understanding (Rossen et al., 2020). Although NPIs have played a critical role in reducing the death toll and number of infected people in the absence of medical treatment, delayed interventions and underestimating the severity of the situation have resulted in over 205 million infected cases, 4.3 million deaths, and billions of dollars in economic losses worldwide (Cutler and Summers, 2020). Due to delayed measures during the initial wave, coronavirus is responsible for between 593,126 and 752,284 excess deaths in the USA (Center for Disease Control, 2021b).

To counteract the economic losses and death toll, the world’s quickest vaccine was invented (Andreadakis et al., 2020). With the proper allocation of the vaccine, the number of infected persons and disease-related mortality has reduced over time, allowing people to return to their normal lifestyles without using NPIs after more than a year (Islam et al., 2021). However, the advent of new variants (delta, lambda, delta plus and so on) is prompting a new pandemic wave to sweep the world right now. With the mass vaccination program, there was an almost exponential drop in the number of infected persons between January 8, 2021 to June 7, 2021 but, with the emergence of new variants, the number of infected people is exponentially growing again.

If proper steps are not implemented quickly, the new variants could produce a considerably worse situation than previous coronavirus waves. The false sense of security can reenact the historic super-spreader incident of the Philadelphia Parade-1918, which resulted in 13000 more deaths in Philadelphia in three months owing to the Spanish flu pandemic (Stetler, 2017).

In light of this situation, in our study, we sought to model scenarios by varying the overall population adoption of NPIs in conjunction with vaccine efficacy, antibody waning, and change in prevalence of the co-circulating variants of the coronavirus over time. One of the most pressing questions during the COVID-19 pandemic was how to correctly parameterize the value of transmission rate (*β*). It has been quite challenging to keep up with the evolution of coronavirus in estimating *β* value. Due to the continual appearance of newer strains of coronavirus with distinct characteristics, estimating a transmissibility value without taking the strains into consideration has now become redundant.

In our model, we have considered a time-varying variant dependent transmission parameter (*β*) by adapting the approach from Islam et al. (2021). To assess the transmissibility, our approach takes into consideration a crucial factor: the changing nature of coronavirus predominant strains over time, making it useful in the present. On the other hand, boosting the number of vaccinated people is one of the most important elements in reducing COVID-19’s devastating effects (Mahmud et al., 2021). As people’s perceptions fluctuate over time, especially with the number of active cases (Center for Disease Control, 2021a), fixed vaccination rate will give a false hope to curb the disease. This method gives a fair picture of how the vaccination rate changes when the number of current cases changes, which is one of the key reasons why individuals become vaccinated (Hamel et al., 2021). Furthermore, with the emergence of new variants, this strategy can be very valuable for planning the third dose and booster dose vaccine based just on historical data of daily cases.

As vaccine protection lasts at least six months (Pfizer Inc, 2021), we observed from our simulations that the best way to reduce number of cases is to increase vaccination rates with high efficacy. However, owing to people’s concern over vaccination safety, NPIs are the only thing that can prevent unvaccinated individuals from falling ill. Interestingly, we found that NPIs had an effect comparable to raising vaccination rates to control the epidemic. As a result, the best method for reducing the spread and death toll is to boost immunization and implement NPIs until herd immunity is achieved. With more data available, this model can provide more accurate projections.

## 2. Methodology

### 2.1. Model Development

We developed an extension of the basic SEIR type compartmental deterministic model to project future disease dynamics in the USA. Our model consists of two distinct tracks for vaccinated and unvaccinated persons, which takes into consideration vaccination rates, waning immunity, viral variations, and reinfection. The compartmental model diagram is shown in figure 1. The model divides the population into seven compartments: unvaccinated susceptible individuals (*S*), vaccinated susceptible individuals (*V*), unvaccinated exposed individuals (*E*), vaccinated exposed individuals (*E*_*v*_), unvaccinated infected individuals (*I*), vaccinated infected individuals (*I*_*v*_), and recovered individuals (*R*).

**Figure 1:**
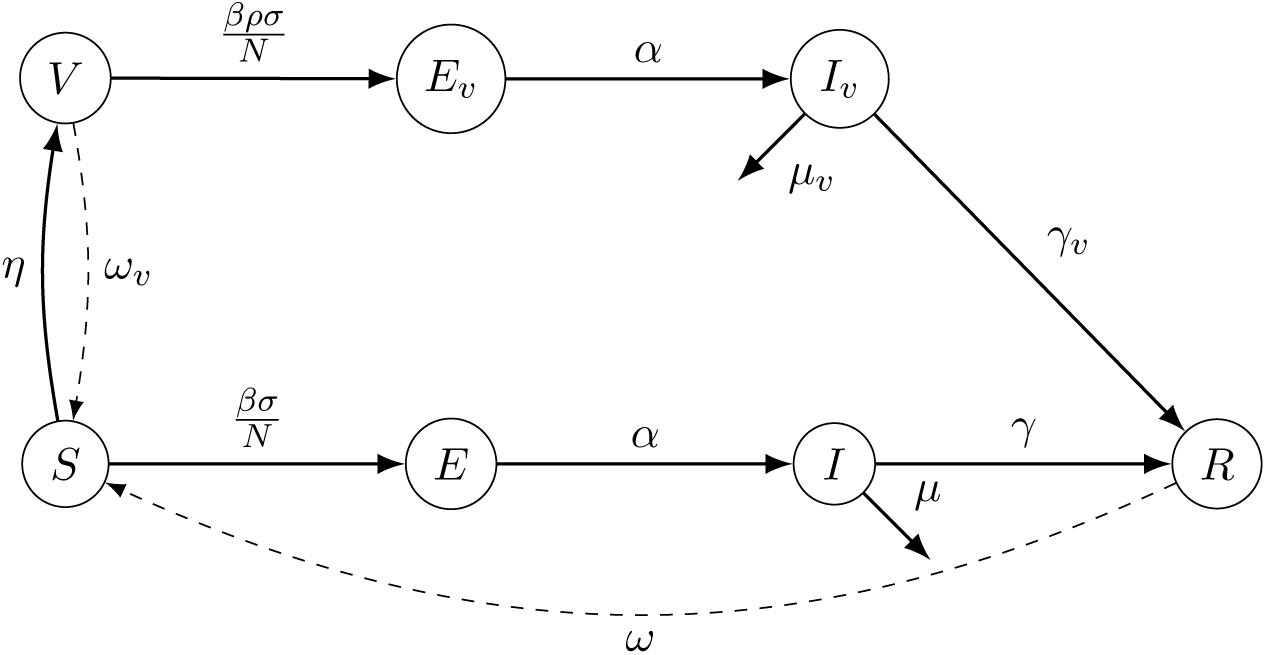
SEIR (Susceptible, Exposed, Infected, Recovered) type Compartmental Model. Here, people moves from susceptible (*S*) and vaccinated (*V*) compartment to exposed compartment (*E* and *E*_*v*_) at 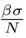 and 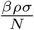 respectively. From *E* and *E*_*v*_ people become infected (*I* and *I*_*v*_) after incubation period *α*. Then from *I* and *I*_*v*_, infected people either go to the recovery compartment *R* or die. With the waning of antibodies, individuals move back to the *S* compartment from the *R* and *V* compartments.

Here, we assume that successful transmission has occurred for both exposed vaccinated and unvaccinated individuals (*E* and *E*_*v*_). Unlike existing models Byambasuren et al. (2020); Buitrago-Garcia et al. (2020); Aguilar et al. (2020); Kronbichler et al. (2020), this model does not explicitly track asymptomatic infections separately from symptomatic infections. This will not impact the result of our model as the asymptomatic cases go to the recovery without getting identified. In fact, there isn’t enough information on asymptomatic cases of the variants. Further to that, due to the increased virulence of the new variants, we focused our research on how the changing proportions of variants affects symptomatic people, for whom the cases may be more deadly than the original strain.

### 2.2. Disease Dynamics

The susceptible population is exposed to the disease at a rate of *β* and vaccinated at a rate of *η*. Although, the vaccinated susceptible population is exposed to the disease at the same rate as the unvaccinated people, but only a fraction of them will become infected due to the vaccine’s protection, that is efficacy of vaccine *ρ*. Once infected, the model assumes that infected individuals have the same level of infectivity regardless of their vaccination status, since viral loads have been found similar between vaccinated and unvaccinated infectious individuals (Acharya et al., 2021).

After the incubation period (*α*), the exposed population goes to *I* and *I*_*v*_ compartment. Then infected persons transfer from *I* and *I*_*v*_ compartments by recovering at *γ* and *γ*_*v*_ or by passing away at *µ* and *µ*_*v*_ rates respectively. We are assuming both vaccinated and non-vaccinated people will take same time to recover. The vaccinated and recovered population again become susceptible owing to the decrease in immunity over time at rates *ω*_*v*_ and *ω* respectively.

The following system of differential equations is our model equations which keep tracks how the individual moves from one compartment to another compartment.

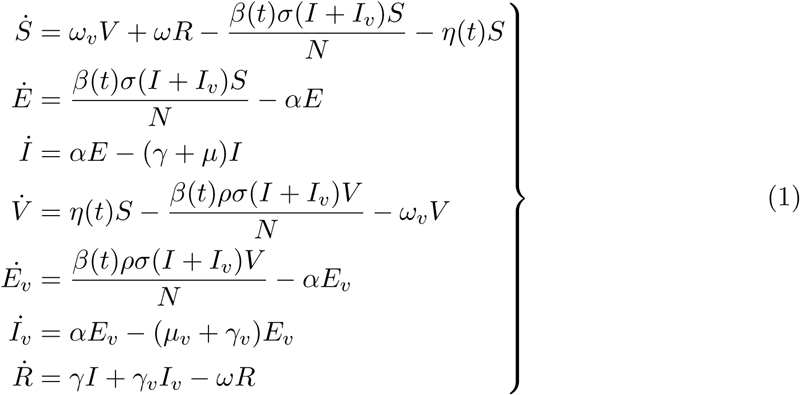

### 2.3. Model Parameterization

Our model consists of ten parameters. Depending on the availability of data, we estimated some and took some of the well established parameters value based on literature. Determining the value of transmissibility, *β*(*t*), is the most challenging parameter of all due to the coronavirus’s ever-changing nature and the abundance of circulating variants. To take into account circulating variants and their relative transmissibility, we used the data from Center for Disease Control (2021a) to generate a weighted average of transmissibility based on relative infectivity and circulating variants prevalence over time by incorporating the approach from Islam et al. (2021). Then *β*(*t*) becomes time dependent that takes into account the emergence of newer virulent variants. To parameterize *β*(*t*), the time dependent relative infectivity, shown in figure 2, is multiplied by a baseline value resulting in ranges from 0.34320 to .5819, which yields effective reproduction numbers in the realistic ranges (Linka et al., 2020; Inglesby, 2020; Jung et al., 2021; Arroyo-Marioli et al., 2021) shown in figure 5.

**Figure 2:**
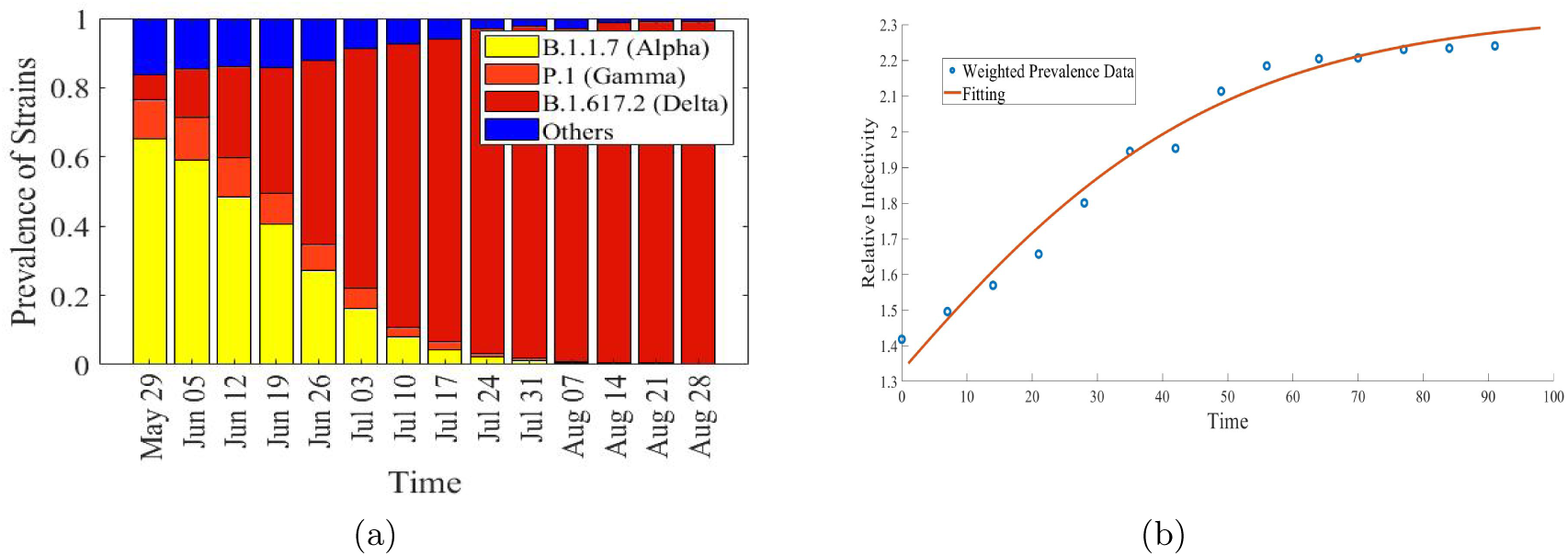
(a) Based on CDC’s Prevalence statistics, the change in prevalence of variants of coronavirus over time from May 29 to August 28 (Center for Disease Control, 2021a). (b) The relative infectivity of circulating viral variants is shown based on 50% increase in infectivity for alpha (B.1.1.7) with respect to other co-circulating variants and a 50% increase in infectivity for delta (B.1.617.2) in comparison to alpha (B.1.1.7) (blue circles). To estimate future relative infectivity, a log-logistic equation is fitted with the model (pink curve) (See Appendix B).

By plotting the daily infection and vaccination data over time from the Center for Disease Control (2021a), we see that the number of people who have been vaccinated per day fluctuates according to the number of infected people per day (figure 3a). We assume that the time-varying vaccination rate (*η*(*t*)) is proportional to the number of active cases based on this phenomenon. If the number of cases rises, more individuals will be interested to take the vaccine. Using the data from Center for Disease Control (2021a), we have calculated the vaccination rate (*η*(*t*)) by using the formula, 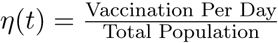, where for total population we have used the most recent value from the U.S. Census Bureau (2020).

**Figure 3:**
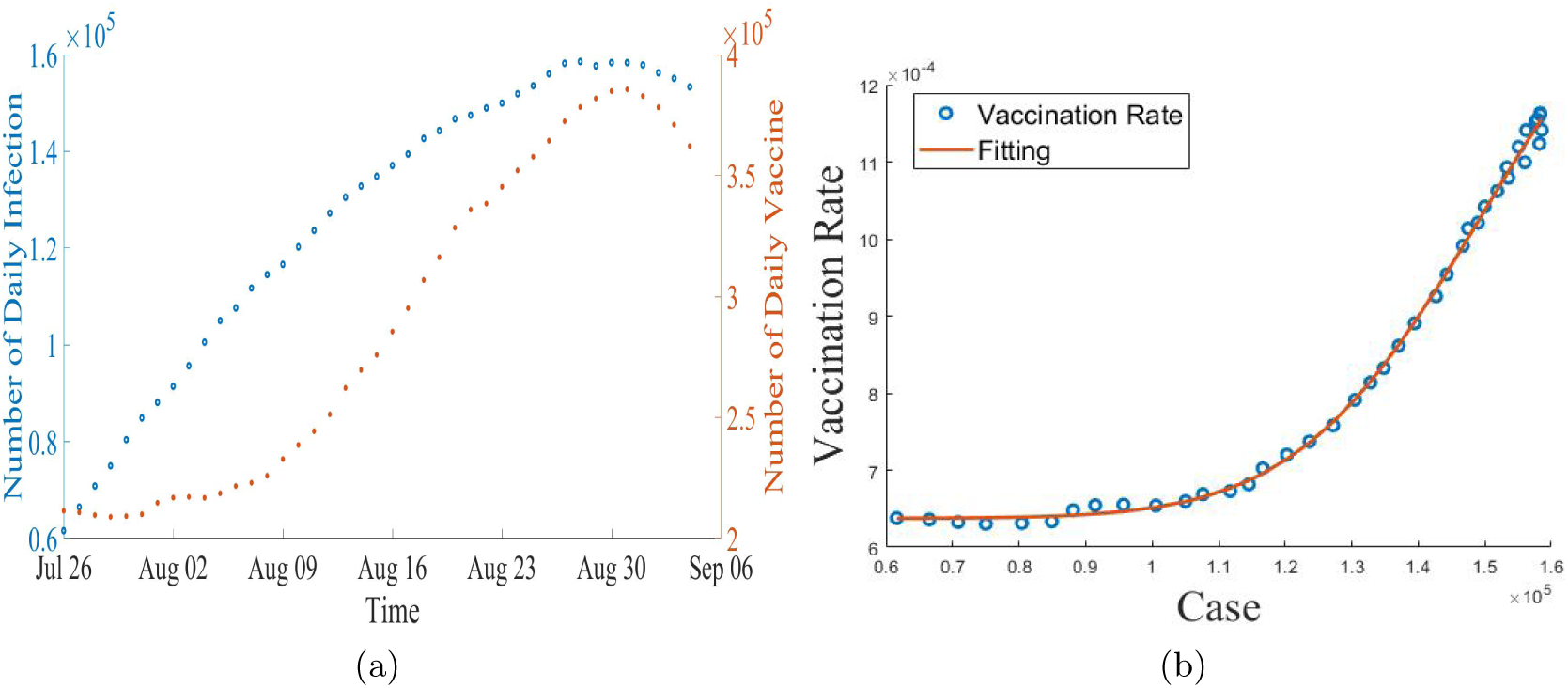
(a) Daily data on infections and vaccination rate from the Center for Disease Control (2021a). (b) The red solid line here represents the log-logistic function which estimates the vaccination rate.

Using the vaccination data from the Center for Disease Control (2021a), we utilized statistical techniques to fit the vaccination rate to the number of daily cases shown in figure 3b. To estimate the vaccination rate using the log-logistic approach, we used the number of daily cases as a predictor to estimate the vaccination rate using the log-logistic function (See Appendix B). This fitted function will predict future vaccination rates based on the values of past cases.

We have assumed the incubation period (*α*) is same for both vaccinated and unvaccinated people to become infected (Center for Disease Control, 2021c). The recovery rate (*γ* and *γ*_*v*_) and disease induced death rate (*µ*) for the non vaccinated persons are adopted from Usherwood et al. (2021). We have used disease induced death rate (*µ*_*v*_) adopted from Vahidy et al. (2021) for vaccinated people.

Based on the recent study, we assumed that the vaccination would be effective for at least 6 months (Pfizer Inc, 2021) and the recovered patients would have the same period of protection as vaccinated persons.

**Table 1:** Model Parameters and Their Description

| Parameter | Value | Source |
| --- | --- | --- |
| $\beta(t)$ (Transmission Rate) | [0.3430 - 0.5819] | Callibrated |
| $\eta(t)$ (Vaccination Rate) | [0.0004 - 0.001] | Estimated |
| $\rho$ (Vaccine Efficacy) | 0.95 | Katella (2021); Pfizer Inc (2021) |
| $\alpha$ (Incubation Period) | $\frac{1}{5} \text{ day}^{-1}$ | Centers for Disease Control and Prevention (2020) |
| $\gamma$ (Recovery Rate) | 0.1 | Usherwood et al. (2021) |
| $\mu$ (Disease Induced Death Rate) | 0.0005 | Usherwood et al. (2021) |
| $\mu_v$ (Vaccinated Disease Induced Death Rate) | 0.000041 | Vahidy et al. (2021) |
| $\omega$ (Antibody Waning rate) | $\frac{1}{180} \text{ day}^{-1}$ | Assumed |
| $\omega_v$ (Vaccine Waning rate) | $\frac{1}{180} \text{ day}^{-1}$ | Pfizer Inc (2021) |
| $\sigma$ (Adoption of NPIs) | [60% - 75%] | Varying the parameter based on Borchering et al. (2021) |

## 3. Results

### 3.1. Analytic Results

The basic reproduction number (*R*_0_) is an epidemiologic statistic used to describe the transmissibility of infectious agents. Although the vaccination of susceptible individuals of the community will limit the number of effective contacts between infectious and susceptible people, this activity will not reduce the *R*_0_ value as *R*_0_ assumes a completely susceptible population (Delamater et al., 2019). As a result, we have estimated the effective reproduction number (*R*_*e*_). The analytic expression of the effective reproduction number (*R*_*e*_) is,

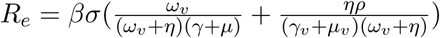

The detailed calculation using the next generation matrix approach of effective reproduction number is shown in the Appendix A. The *R*_*e*_ value, rather than the *R*_0_ value, would be reduced if the number of susceptible people in a population were reduced through vaccination. If *R*_*e*_ can be decreased to a value *<* 1, immunization might potentially halt an epidemic (Anderson and May, 1992; Anderson, 1992; Rubió, 2012).

### 3.2. Co-Circulating Variants, Vaccine Efficacy and NPIs

Keeping a steady rate of viral transmission, we simulated the scenarios based on the adoption of NPIs and vaccine effectiveness (figure: 4). The reduction in vaccine efficacy occurs with time as well as with the advent of new vaccine breakthrough variants. People, on the other hand, are adopting NPIs as they see fit because there is no set of mandates for individuals to follow when it comes to NPI strategy, making it impossible to get information on which NPI approach is employed by how many people. Instead of concentrating on which NPIs are used, it is now more appropriate to take into account the proportion of the population that uses NPIs as a measure of prevention.

**Figure 4:**
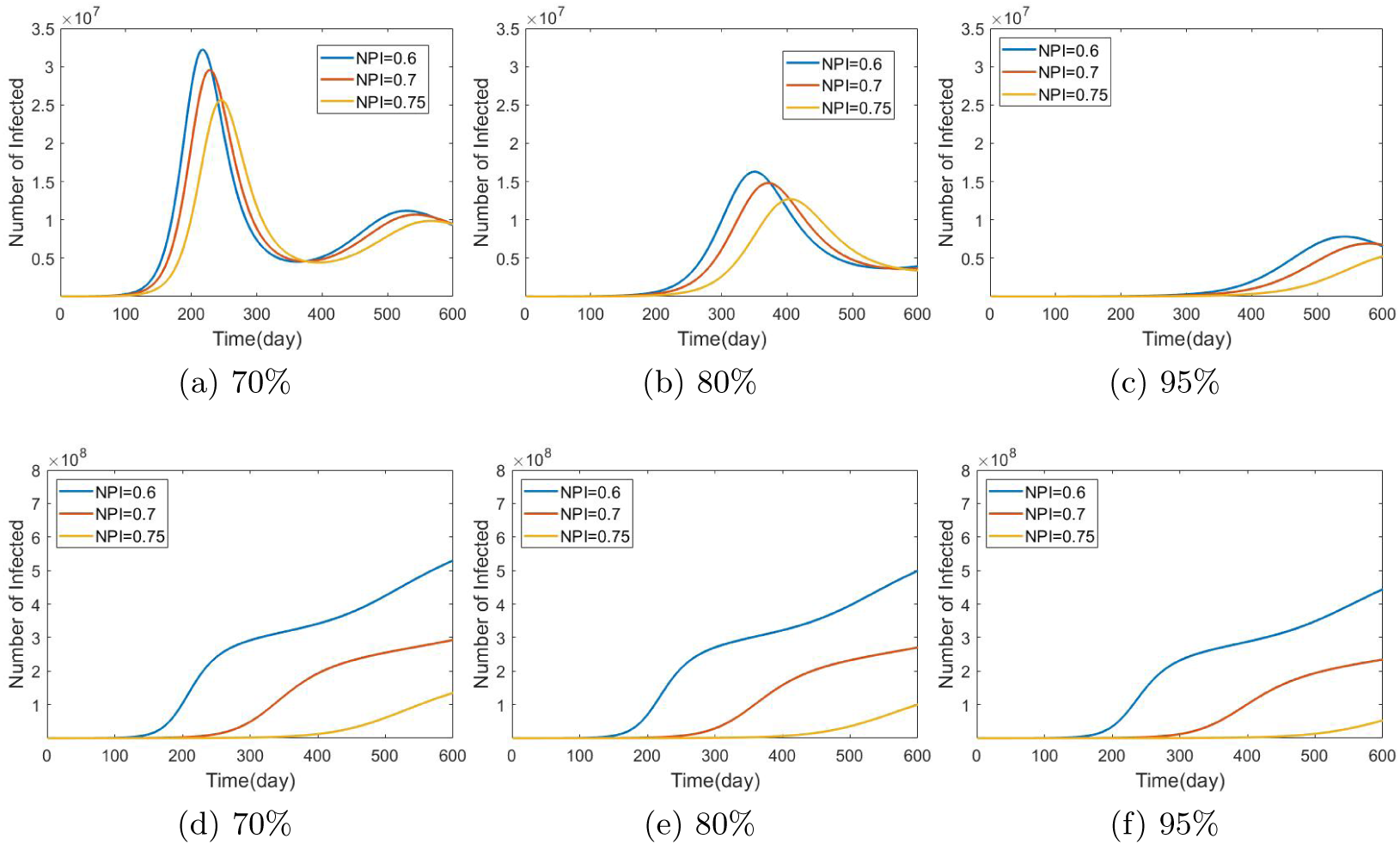
The effect of NPIs adoption and vaccine efficacy on the daily number of infected cases is depicted in this figure. Figures a, b, and c depict the change in daily case count with fixed vaccine efficacy (70%, 80%, 95%) with changing adoption of NPIs, while figures d, e, and f depict the corresponding cumulative case count. From a, b, c, d, e, f it is clear that if the majority of the population uses NPIs, NPIs will have the greatest influence on lowering the daily number of instances in the short and long term. If NPI adoption is not achievable, the only way to minimize the number of cases is to have a high vaccination rate with high effectiveness. (Check out Appendix C for a comparison of peak values.)

To simulate the scenarios of waning efficacy of vaccine, we varied the effectiveness of the vaccine from 70% to 95% (figure 4a, 4b, 4c). In the simulation, as the efficacy rises, the number of persons infected decreases significantly, implying that a highly effective vaccine is needed to reduce the number of infections in the short and long term.

On the other hand, if the number of infected people decreases, people will be less likely to use NPIs which is why we have varied the total populations adoption of NPIs to simulate the situations with different adoption rate. Figure 4 indicates that even if 75% of the population uses NPIs, the number of sick people will remain relatively low, regardless of vaccine effectiveness. It implies that using NPIs, until herd immunity is obtained or high vaccine efficacy is maintained indefinitely, is the best strategy.

### 3.3. Effective Reproduction Number

The effective reproduction number, *R*_*e*_, is affected by changes in immunization rate, vaccine efficacy, and NPI adoption. While vaccine efficacy cannot be modified quickly, NPI adoption and vaccination rate can change drastically. Also, against the new variants, vaccine loses its efficacy (Center for Disease Control, 2021d) for which we calculated the *R*_*e*_ value by setting the vaccine effectiveness to 80% instead of 90% while varying the vaccination rate, *η*(*t*) from 0.0001 to 0.003, as well as the NPIs adoption rate from 0% to 75%. The effective reproduction number fluctuates between 4.31 (worst scenario) and 0.78 (best scenario)(figure: 5).

**Figure 5:**
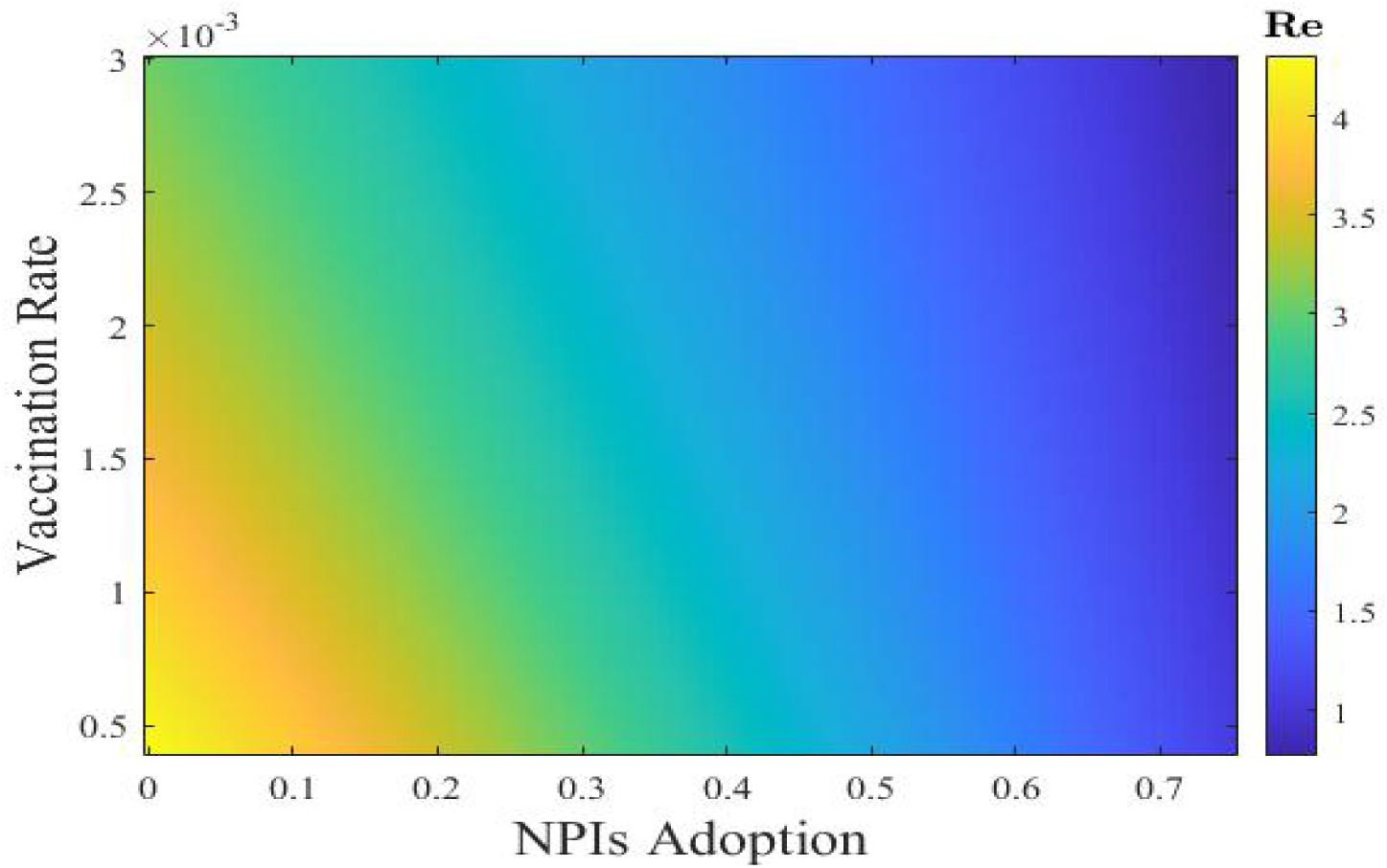
Heat-map here depicts how with fixed vaccine efficiency the effective reproduction number (*R*_*e*_) varies with vaccination rates and adoption of NPIs. With vaccine efficacy 80%, the most efficient method to halt the spread of the disease is to increase immunization rates paired with good implementation of NPIs until herd immunity is reached.

**Figure 6:**
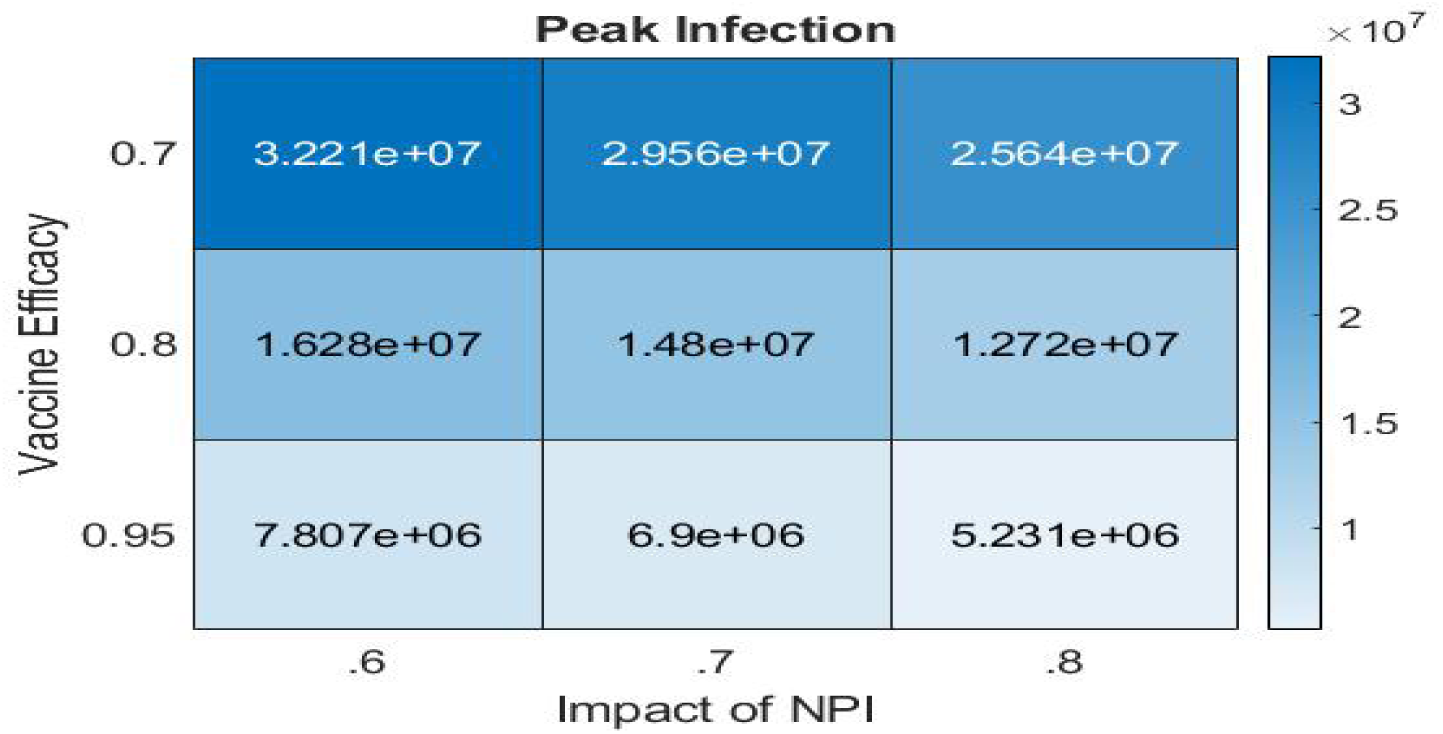
The variation in the peak value of daily infected cases as a function of NPI uptake and vaccination effectiveness. With an increase in NPI uptake or vaccine efficacy, the peak value is much lower.

## 4. Discussion

The adoption of NPIs, effective allocation, and deployment by assuring the high efficacy of the vaccine against the virus throughout time are the most significant components in controlling the spread of the COVID-19. With the introduction of novel coronavirus variants and declining immunity over time, along with an increasing perception of safety, has produced an ideal environment for the superspreading of highly infectious and lethal coronavirus variants. In this situation, we developed an extension of the standard compartmental SEIR type model that dynamically tracks changes in co-circulating variants and dynamic vaccination rates. As time passes, the emergence of new coronavirus variants renders a fixed transmission rate obsolete, because each variant differs in terms of transmission potential. The overall coronavirus transmission rate changes over time depending on the transmissibility of the co-circulating variants. As a result, tracking the change in *β*(*t*) value with the change in co-circulating variants over time is a useful technique to track the transmission rate of coronavirus at any given time. Furthermore, the vaccination data was not uniform at first due to people’s anxiety and misinformation about the COVID-19 vaccine. Based on our data fitting, it is clear that the vaccination rate currently follows the trend in daily number of cases with a lag, allowing us to create a time-varying case-dependent vaccination rate. This strategy particularly accounts for people’s shifting attitudes toward vaccination as the number of cases grows, making it a practical way to account for variants and vaccination rates in the current situation.

To reduce the complexity caused by the continual change in variants, we did not explicitly include asymptomatic infected people in our model, instead focusing on the shifting landscape of variant strains and parameterizing relative transmissibility using empirical time series weighted prevalence data for symptomatic people (figure 2). Furthermore, rather than indicating which NPIs are most beneficial, we now state what percentage of the population has implemented NPIs. This is the logical thing to do right now, because individuals are no longer adopting a predetermined set of NPIs plan, but rather choosing to protect themselves against the COVID-19 in their own way, as states no longer have a mandate. Moreover, to avoid the ambiguity about the efficacy of different NPIs measures, we concentrated on the overall adoption of NPIs by the proportion of the population.

Based on the findings of our model, we can definitely claim that vaccination rate, vaccine effectiveness, and NPIs are significant factors in reducing the number of infected patients and controlling the disease. According to our simulation, if vaccine efficacy declines slightly, the number of sick people increases considerably, demonstrating that vaccine efficacy is crucial for viral containment. As a result, the race to create and deliver booster dose and third dose is critical right now in order to maintain the vaccine’s efficacy against new strains. On the other hand, if mass vaccination is not possible in a timely way or if vaccine breakthrough variants become prevalent, NPIs are the only option for controlling the spread of COVID-19. The project’s next phase will be to add asymptomatic individuals with strain-based data. Furthermore, vaccination data will grow more consistent over time, improving the accuracy of this model. Therefore, this modeling technique highlights the importance of the individuals to become vaccinated, as well as the need of planning and distributing booster dose in a timely manner in order to mitigate the spread of coronavirus.

## Data Availability

All data produced in the present study are available upon reasonable request to the authors.

## Acknowledgement

We would like to thank Dr. Linda J.S. Allen, Professor, Department of Mathematics and Statistics, Texas Tech University, for suggestions with the project.

## Supplementary Materials

### A. Effective Reproduction Number

The Jacobian matrix J is obtained from linearizing the system (1). The disease-free equilibria of the model at *X*_0_ = (*S*_0_, 0, 0, *V*, 0, 0, 0); where 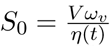 is the initial population of susceptible individuals. Now, reorganizing the DFE using, *N* = *S* + *V* as at DFE the population are either in susceptible compartment or in vaccinated compartment, we get, 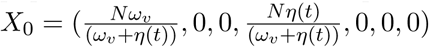. Evaluating the Jacobian matrix at *X*_0_ yields,

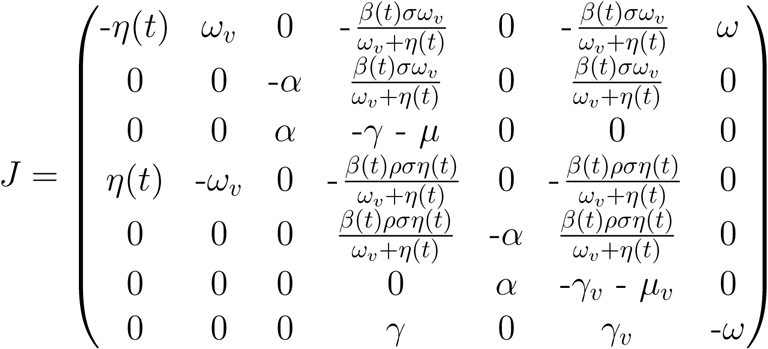

The linearized infected subsystem of (1) evolves according to the following set of equations for minor perturbations *z* = (*S, E, I, V, E*_*v*_, *I*_*v*_, *R*) near the *X*_0_

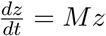

Here,

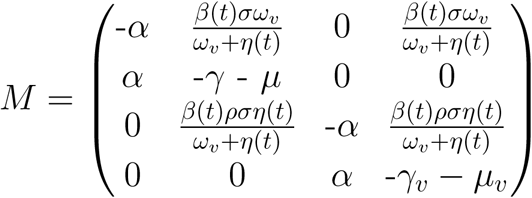

We decompose the matrix M into transmission (T) and transition (∑) matrices respectively, obtaining

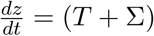

where

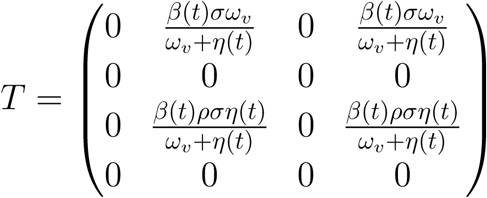

and

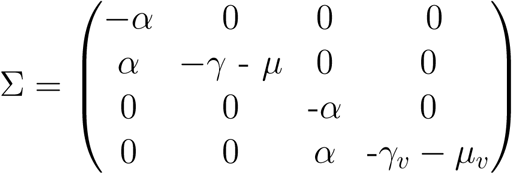

K is a large-domain next-generation matrix. Because T is ranked first, the NGM K is likewise ranked first. As a result, non-zero entries are only found in the first row of K. Therefore, K’s spectral radius is the first item on the diagonal, i.e., *K*_11_, which is identical to *R*_*e*_. Therefore,

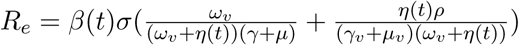

### B. Data Fitting

#### B.1. Relative Infectivity

Relative infectivity is fitted against the number of days as time from May 29, 2021 to August 28, 2021 using the following logistic model.

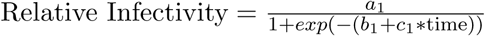

where as the p-values are within range, all of the parameters *a*_1_, *b*_1_ and *c*_1_ are statistically significant. The fitted model:

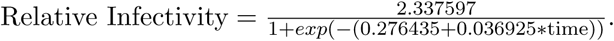

**Table 2:** Parameter Estimates for Relative Infectivity

| Parameters | Estimates | t-values | p-values |
| --- | --- | --- | --- |
| $a_1$ | 2.337598 | 50.577 | $2.22e^{-14}$ |
| $b_1$ | 0.276434 | 4.906 | 0.000467 |
| $c_1$ | 0.036925 | 8.654 | $3.07e^{-06}$ |

#### B.2. Vaccination Rate

Based on the daily number of cases, we built a log-logistic model to estimate the vaccination rate. The fitted model is

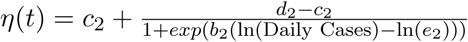

where all the estimate coefficient of the daily instances is statistically significant as all the *p*-values are well below 0.05.

The fitted model:

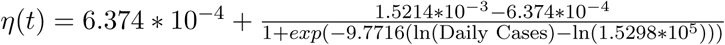

### C. Peak Infection

The number of infections at their peak fluctuates dramatically depending on the circumstance. This statistic is critical for the health sector to take adequate precautions in order to prepare for the worst-case scenario in terms of affected persons.

